# Widespread Programs, Limited Evidence: How U.S. MD-Granting Medical Schools Define, Measure, and Evaluate Student Well-Being Efforts

**DOI:** 10.64898/2026.10.21.26366188

**Authors:** Jayson Guo, Maia Winkel, Al’ai Alvarez, Carl Preiksaitis

**Affiliations:** John A. Burns School of Medicine, University of Hawai’i at Mānoa, Honolulu, HI, USA; Department of Emergency Medicine, Stanford University School of Medicine, Stanford, CA, USA

**Keywords:** Students, Medical, Curriculum, Health Promotion, Program Evaluation, Surveys and Questionnaires

## Abstract

**Purpose:** To characterize how U.S. MD-granting medical schools define wellness aims, assess student well-being, evaluate program effectiveness, and analyze and report outcomes.

**Method:** From June 2025 through September 2026, the authors surveyed all 159 Liaison Committee on Medical Education-accredited U.S. MD-granting schools using a web-based instrument. Analyses used item-specific denominators for partial responses. Among 84 schools answering five linked items, the authors examined completion of the sequence from defined aims to reported outcomes. Exploratory analyses examined associations between explicit wellness competencies and the remaining practices.

**Results:** Of 159 schools, 107 (67.3%) provided substantive responses. Of the 84 schools that answered all five linked items, 2 (2.4%) reported the complete sequence, and 22 (26.2%) reported none. Explicit wellness competencies were reported by 23/91 schools (25.3%); 38/91 (41.8%) were unsure whether any existed. Schools reported formal assessment of student well-being (48/91, 52.7%) and formal program evaluation (43/90, 47.8%). Among schools that evaluated, 39/42 (92.9%) used Association of American Medical Colleges national surveys, whereas 14/42 (33.3%) used any standardized longitudinal or pre/post measure. Schools rarely analyzed outcomes by student demographic characteristics (16/89, 18.0%), and none reported publishing outcomes in full. Schools with explicit wellness competencies reported more of the remaining practices (median 2 vs 1, *P* = .007), with associations concentrated in program evaluation and outcome reporting. Wellness infrastructure was common: dedicated curricula (65/107, 60.7%), learning communities (85/98, 86.7%), mistreatment reporting or prevention policies in clinical training (89/97, 91.8%), and wellness committees (73/84, 86.9%).

**Conclusions:** Responding U.S. medical schools commonly reported wellness infrastructure, but few reported a complete sequence from defined wellness aims to reported outcomes. Gaps in explicit wellness competencies, assessment, program evaluation, and outcome reporting limit schools’ ability to judge program effectiveness. Explicit wellness competencies were associated with more evaluation and reporting practices; whether they improve program effectiveness remains unclear.

## Introduction

Medical students enter training with lower rates of burnout and depressive symptoms than similarly aged college graduates,^1^ yet approximately half of students experience burnout during training and more than one quarter report depressive symptoms.^2,3^ Shortly before graduation, 49% of students in one multi-institutional sample had burnout and 38% endorsed depressive symptoms.^4^Although these studies did not follow the same students, they suggest that training conditions contribute to declining well-being. Student distress is associated with poorer academic performance, reduced empathy, unprofessional behavior, substance misuse, and consideration of withdrawal from training.^5^ This distress reflects both individual vulnerabilities and the conditions of medical education, including demanding curricula, high-stakes assessment, mistreatment, and inadequate support.^5-7^ Medical student well-being is therefore both an individual health concern and an institutional responsibility. The Liaison Committee on Medical Education (LCME) requires effective counseling and programs that promote student well-being.^8^

Medical schools have expanded their wellness initiatives, but two prior multi-institutional surveys identified gaps in program evaluation. Dyrbye and colleagues surveyed 32 U.S. medical schools in October 2016; 27 (84%) responded. Wellness curricula and learning communities were widespread, but only 6 of 27 schools (22%) reported explicit wellness competencies, and program evaluation relied largely on participation rates and satisfaction ratings.^9^ Schutt and colleagues surveyed 159 U.S. and Canadian schools in July 2019; 104 (65%) responded. Formal wellness programs were nearly universal, but structural programming was often considered insufficient, and program evaluation relied heavily on the Association of American Medical Colleges (AAMC) Graduation Questionnaire.^10^ An AAMC working group noted that national survey instruments contain little wellness-specific content and recommended pairing these instruments with brief standardized measures aligned with local program goals.^11^

These studies establish that wellness programs are common but leave a central question unanswered: whether schools can determine what programs accomplish. The Centers for Disease Control and Prevention (CDC) Program Evaluation Framework provides a structure for answering that question.^12^ The framework calls for describing a program’s intended outcomes, gathering credible evidence, generating conclusions, and acting on the findings, with advancing equity and collaborative engagement as cross-cutting commitments. Applied to institutional wellness efforts, these steps suggest five linked practices: defining wellness aims, assessing student well-being with measures that can detect change, evaluating whether programs achieve those aims, analyzing outcomes by student demographic characteristics, and reporting findings to those who can act on them. Gaps in this sequence limit schools’ ability to interpret outcomes and use findings to improve programs. Prior surveys examined several practices individually but did not assess whether schools linked them. No survey has reassessed the U.S. landscape since 2019, before USMLE Step 1 transitioned to pass/fail reporting and digital wellness tools expanded.

We therefore surveyed all 159 LCME-accredited U.S. MD-granting schools. Our aims were to update national estimates of wellness programming and organizational support, and to characterize five practices: defining wellness aims, assessing student well-being, evaluating wellness programs, analyzing outcomes by student demographic characteristics, and reporting results.

## Method

### Survey development

Drawing on prior national surveys and related literature on medical student well-being,^9-11^we developed a cross-sectional institutional survey covering four domains: wellness curricula and structural features of the learning environment; assessment of student well-being and evaluation of programs; organizational and operational resources; and future priorities. The survey included 36 single-response, multiple-response, or open-text items. Item-level denominators varied because some questions were optional and display logic presented follow-up items only when applicable.

Content and response-process validity evidence^13^ came from two physicians with well-being expertise (A.A., M.W.): a department well-being director who leads a physician wellness fellowship and directs the physician peer support program for a large academic health system, and a current physician wellness fellow. Both reviewed item content and completed the Qualtrics survey in think-aloud cognitive interviews, verbalizing their interpretation of each item. Their feedback informed revisions to item wording, response options, and content coverage, including clarifying the explicit wellness competencies item and adding an item assessing follow-up support after the wellness assessment. Pilot responses were excluded from the analytic dataset. The complete instrument is provided in Supplemental Digital Appendix 1.

### Sampling and recruitment

The sampling frame included all 159 LCME-accredited U.S. MD-granting medical schools as of May 2025.^14^ For each school, we searched publicly available websites to identify a key informant. We prioritized a well-being director or dean; otherwise, we selected a student-affairs leader whose role or biography indicated responsibility for student well-being. When available, we recorded additional contacts and occasionally telephoned student-affairs offices to identify the most appropriate respondent.

We administered the survey in Qualtrics from June 26 through October 28, 2025, and conducted a supplementary round from August 4 through September 15, 2026, for schools that had not responded. We sent each contact a personalized link, requested one response per school, and encouraged recipients to forward the invitation internally or identify a more appropriate colleague. The landing page stated that completion would take 15 to 20 minutes and that school-level responses would remain confidential and be reported in aggregate. Respondents could also complete the survey by telephone, with a study investigator entering answers. We sent automated reminders at approximately weekly intervals, and repeated personalized outreach to secondary and tertiary contacts across sequential distribution rounds. We offered no incentive.

### Data management and analysis

Because outreach reached multiple contacts per school and one school received a reissued survey link, the export contained more records than schools. We mapped each record to its school and analyzed the data at the school level, retaining the most complete record when a school had more than one. For all but one affected school, the discarded record contained no substantive answers; for the remaining school, two substantive records were present, and the less complete was excluded. We then excluded schools whose only records contained no answer to any substantive item. We did not impute missing values. Counts and percentages used item-specific available-case denominators, follow-up denominators were restricted to respondents eligible through display logic, and multiple-response items were summarized as the number of schools selecting each option.

Five items, although not designed from the CDC framework, corresponded to its steps and crosscutting actions and defined the sequence from stated aims to reported evidence: explicit wellness competencies (describing the program), formal assessment of student well-being (gathering credible evidence), formal evaluation of program effectiveness (generating conclusions), analysis of outcomes by student demographic characteristics (advancing equity), and any reporting of summary outcomes beyond the responsible office (acting on findings). This analysis was restricted to schools answering all five. As an exploratory analysis, we examined whether schools reporting explicit competencies differed from all others in the number of the remaining four practices reported, using a Mann-Whitney U test as the single primary comparison. Associations with each individual practice and with two organizational supports are reported as odds ratios with exact 95% confidence intervals and Holm-adjusted Fisher exact *P* values (Supplemental Digital Appendix 2). Because respondents unsure about competencies may also have been less familiar with other practices, we repeated these comparisons excluding them.

We assessed potential nonresponse bias by comparing responding and nonresponding schools across the full sampling frame by AAMC region using a chi-square test, and by initial accreditation before 2000 versus 2000 or later, and by full accreditation without probation versus another status, using Fisher exact tests and standardized differences. To assess whether declining item denominators reflected differential attrition, we compared schools that completed the full instrument with those that provided partial responses on items both groups reached. Analyses were conducted in Python version 3.11 with SciPy version 1.17 (Python Software Foundation, Wilmington, Delaware).^15^ The authors used Claude (Anthropic) to help write analysis code and edit text, and verified all code, results, and text. Reporting followed the Checklist for Reporting of Survey Studies and Academic Medicine guidance for survey-based research.^16,17^

One investigator reviewed substantive responses to selected open-text prompts, identified recurring content, and applied the resulting nonexclusive categories through keyword search and manual review. Categories are sometimes nested, with the surgery clerkship counted within clinical rotations and Step 1 within board examinations. Counts are reported to contextualize closed-ended findings; we did not conduct formal qualitative thematic analysis or assess intercoder agreement, and these counts should not be read as national prevalence estimates.

### Ethical review

On March 26, 2025, the Stanford University Institutional Review Board determined that the project did not involve human subjects as defined in 45 CFR 46.102(e) or 21 CFR 50.3(g) (protocol 79558); no further action was required.

## Results

### Participation

After deduplication and excluding records with no substantive answer, 107 schools remained for analysis, representing 67.3% of the 159 invited schools; 16 responded during the supplementary round. Eighty-three schools reached the end of the survey, and 24 stopped partway; because items were optional, denominators fell from 107 for the first item to 81 for the last closed-ended item.

Response rates were 80.0% (28/35) in the Central region, 75.0% (18/24) in the Western region, 63.8% (37/58) in the Southern region, and 57.1% (24/42) in the Northeast (*χ*^2^ = 5.50, *P* = .14, Cramér *V* = 0.19). Response did not differ by initial accreditation era (2000 or later, 24/35 [68.6%], vs before 2000, 83/124 [66.9%]; standardized difference 0.04; *P* > .99). Schools holding full accreditation without probation responded somewhat more often than others (101/148 [68.2%] vs 6/11 [54.5%]; standardized difference +0.28; *P* = .34), a comparison involving only 11 schools. Schools completing the full instrument did not differ from partial responders on items both groups reached, including dedicated wellness curricula (52/83 [63%] vs 13/24 [54%]; *P* = .48), learning communities (73/83 [88%] vs 12/15 [80%]; *P* = .41), and formal assessment of student well-being (44/83 [53%] vs 4/8 [50%]; *P* > .99), suggesting that declining denominators did not systematically favor schools with more wellness activity.

### Wellness programming and the learning environment

Sixty-five of 107 schools (60.7%) reported a dedicated wellness curriculum, and 13 (12.1%) reported one in development. Among 75 schools describing its structure, wellness content was most often part of orientation programming (62/75, 82.7%), followed by standalone courses or workshops (54/75, 72.0%) and integration into existing coursework or rotations (48/75, 64.0%). Curricula targeted preclinical students (70/75, 93.3%) and clinical students (62/75, 82.7%; Table 1).

**Table 1.** Wellness Curricula, Learning-Environment Strategies, and Organizational Support Reported by 107 U.S. MD-Granting Medical Schools, 2025-2026.

| Survey item and response | No./total (%) |
| --- | --- |
| **Dedicated wellness curriculum** |  |
| Yes | 65/107 (60.7) |
| In development or actively planning | 13/107 (12.1) |
| No | 29/107 (27.1) |
| **Curriculum structure (multiple responses)** |  |
| Part of orientation programming | 62/75 (82.7) |
| Standalone courses or workshops | 54/75 (72.0) |
| Integrated into existing coursework or rotations | 48/75 (64.0) |
| **Curriculum audience (multiple responses)** |  |
| Preclinical students | 70/75 (93.3) |
| Clinical students | 62/75 (82.7) |
| **Clinical learning-environment strategies (multiple responses)** |  |
| Mistreatment reporting or prevention policies | 89/97 (91.8) |
| Work-hour limits or protected time | 69/97 (71.1) |
| Grading or assessment modifications intended to reduce stress | 64/97 (66.0) |
| Wellness or reflection check-ins | 56/97 (57.7) |

Table 1 (continued)
| Survey item and response | No./total (%) |
| --- | --- |
| Training or guidance for faculty and residents | 46/97 (47.4) |
| Flexible scheduling | 32/97 (33.0) |
| Healthy food or guaranteed meal breaks | 19/97 (19.6) |
| **Faculty development (multiple responses)** |  |
| Training to enhance faculty well-being | 53/98 (54.1) |
| Training to help faculty support student wellness | 43/98 (43.9) |
| **Learning communities** |  |
| Structured learning communities offered | 85/98 (86.7) |
| Include faculty | 79/84 (94.0) |
| Include wellness, self-care, or resilience programming | 76/84 (90.5) |
| Include students from different years | 67/84 (79.8) |
| **Organizational support** |  |
| Designated administrative wellness leader | 65/84 (77.4) |
| Student wellness committee (any composition) | 73/84 (86.9) |
| Dedicated institutional budget line | 63/82 (76.8) |
| **Student participation beyond committee membership (multiple responses)** |  |
| Leading student wellness organizations | 70/82 (85.4) |
| Serving in designated wellness leadership roles | 52/82 (63.4) |
| Conducting student-led research or quality improvement | 48/82 (58.5) |
| Co-designing wellness curricula or programming | 45/82 (54.9) |
| **Barriers to curricular or structural change (multiple responses)** |  |
| Curricular overcrowding | 75/91 (82.4) |
| Logistical or scheduling difficulties | 57/91 (62.6) |
| Lack of financial resources | 40/91 (44.0) |
| Limited administrative or leadership support | 20/91 (22.0) |
| Faculty resistance or lack of buy-in | 19/91 (20.9) |

Note: Denominators vary because partial responses were retained and display logic limited some follow-up items. Percentages for multiple-response items sum to more than 100%.

Structured learning communities were reported by 85 of 98 schools (86.7%); among the 84 describing their content, 76 (90.5%) included wellness, self-care, or resilience programming and 79 (94.0%) included faculty. In the clinical learning environment, mistreatment reporting or prevention policies were near-universal (89/97, 91.8%), followed by work-hour limits or protected time (69/97, 71.1%) and grading or assessment modifications intended to reduce stress (64/97, 66.0%). Less common were training or guidance for faculty and residents on promoting wellness (46/97, 47.4%) and flexible scheduling (32/97, 33.0%). On a separate faculty-development item, 53 of 98 schools (54.1%) offered training to enhance faculty well-being and 43 (43.9%) offered training to help faculty support student wellness.

Of the 52 schools that described notable curricular strategies in open text, 19 (36.5%) named learning communities or similar longitudinal mentoring structures. Among 63 describing approaches they considered effective, access to counseling or mental health services was most common (26/63, 41.3%), followed by student involvement in wellness work (23/63, 36.5%).

Curricular overcrowding was the most frequently reported barrier to curricular or structural change (75/91, 82.4%), followed by logistical or scheduling difficulties (57/91, 62.6%) and limited finances (40/91, 44.0%). Among 64 schools naming particularly stressful elements of their own curriculum, the most common categories were curricular workload or content (46/64, 71.9%), clinical clerkships or rotations (39/64, 60.9%), and board or licensing examinations (32/64, 50.0%); the surgery clerkship (15/64, 23.4%) and Step 1 specifically (19/64, 29.7%) were each named by about one quarter.

### Organizational support and student participation

Sixty-five of 84 schools (77.4%) reported a designated administrator whose primary responsibility included student wellness, and 63 of 82 schools (76.8%) reported a dedicated institutional budget line.

Seventy-three of 84 schools (86.9%) reported wellness committees, most often as student-faculty collaborations (47/84, 56.0%) or predominantly student bodies (24/84, 28.6%). Committee functions extended beyond event planning (72/73, 98.6%) to peer support and outreach (59/73, 80.8%), liaison with faculty and administration (53/73, 72.6%), policy advocacy (44/73, 60.3%), and management of a dedicated budget (31/73, 42.5%). Beyond committees, 70 of 82 schools (85.4%) reported students leading wellness organizations, 52 (63.4%) reported designated student wellness leadership roles, and 45 (54.9%) reported student participation in co-designing curricula or programming.

### Defined aims

Twenty-three of 91 schools (25.3%) reported explicit wellness competencies. Thirty (33.0%) reported none, and 38 (41.8%) were unsure whether their school had any, the highest proportion of uncertainty for any item in the survey (Table 2).

**Table 2.** Defined Aims, Measurement of Student Well-Being, Program Evaluation, and Use of Wellness Data Reported by 107 U.S. MD-Granting Medical Schools, 2025-2026.

| Survey item and response | No./total (%) |
| --- | --- |
| **Explicit wellness competencies** |  |
| Yes | 23/91 (25.3) |
| No | 30/91 (33.0) |
| Unsure | 38/91 (41.8) |
| **Formal assessment of student well-being** |  |
| Yes | 48/91 (52.7) |
| No | 32/91 (35.2) |
| Unsure | 11/91 (12.1) |
| **Assessment methods among assessing schools (multiple responses)** |  |
| AAMC national surveys | 44/48 (91.7) |
| Local attitudinal or experience survey | 30/48 (62.5) |
| Focus groups, town halls, or interviews | 25/48 (52.1) |
| Other standardized well-being instrument | 24/48 (50.0) |
| Standardized burnout instrument | 13/48 (27.1) |

Table 2 (continued)
| Survey item and response | No./total (%) |
| --- | --- |
| Standardized depression instrument | 12/48 (25.0) |
| Standardized anxiety instrument | 11/48 (22.9) |
| Any standardized instrument | 28/48 (58.3) |
| AAMC surveys without any standardized instrument | 18/48 (37.5) |
| **Follow-up after assessment** |  |
| Opt-in support (student must request) | 25/48 (52.1) |
| Opt-out support when defined criteria met | 8/48 (16.7) |
| No follow-up support | 9/48 (18.8) |
| Unsure | 6/48 (12.5) |
| **Anonymous self-administered screening** |  |
| Offered | 44/90 (48.9) |
| **Formal evaluation of program effectiveness** |  |
| Yes | 43/90 (47.8) |
| No | 31/90 (34.4) |
| Unsure | 16/90 (17.8) |
| **Evaluation methods among evaluating schools (multiple responses)** |  |
| AAMC national surveys | 39/42 (92.9) |
| Wellness-specific satisfaction ratings | 35/42 (83.3) |
| Attendance or participation rates | 29/42 (69.0) |
| Qualitative feedback | 27/42 (64.3) |
| Longitudinal tracking with standardized instruments | 11/42 (26.2) |
| Standardized pre/post assessment | 10/42 (23.8) |
| Any standardized longitudinal or pre/post measure | 14/42 (33.3) |
| **Barriers to program evaluation (multiple responses)** |  |
| Limited staffing or personnel time | 45/84 (53.6) |
| Low student participation or engagement | 36/84 (42.9) |
| Insufficient financial resources | 29/84 (34.5) |
| Lack of suitable instruments | 22/84 (26.2) |
| Lack of expertise | 7/84 (8.3) |
| **Analysis of outcomes by student demographic characteristics** |  |
| Yes | 16/89 (18.0) |
| No | 50/89 (56.2) |
| Unsure | 23/89 (25.8) |
| **Publication of summary wellness outcomes** |  |
| Fully | 0/84 (0.0) |
| Partially | 9/84 (10.7) |
| No | 57/84 (67.9) |
| Unsure | 18/84 (21.4) |

Abbreviation: AAMC, Association of American Medical Colleges. Note: Denominators vary because partial responses were retained and display logic limited some follow-up items. Percentages for multiple-response items sum to more than 100%. Standardized instruments are published instruments with validity evidence for the construct measured (eg, Maslach Burnout Inventory, PHQ-9); the survey item labeled these "standardized, validated instruments."

### Measurement of student well-being

Forty-eight of 91 schools (52.7%) formally assessed student well-being; 32 (35.2%) did not, and 11 (12.1%) were unsure. Among the 48 assessing schools, AAMC national surveys were nearly universal (44/48, 91.7%), followed by local attitudinal or experience surveys (30/48, 62.5%), focus groups or town halls (25/48, 52.1%), and other standardized well-being instruments with published validity evidence, such as the Medical Student Well-Being Index (24/48, 50.0%). Condition-specific standardized instruments were less common: burnout (13/48, 27.1%), depression (12/48, 25.0%), and anxiety (11/48, 22.9%). Taken together, 28 of 48 assessing schools (58.3%) used at least one standardized instrument, whereas 18 (37.5%) used AAMC surveys without any standardized instrument.

Follow-up after assessment was most often passive. Twenty-five of 48 schools (52.1%) offered optin support that required students to request services, 8 (16.7%) automatically offered support to students who met defined criteria, 9 (18.8%) provided no follow-up support, and 6 (12.5%) were unsure. Anonymous self-administered screening tools were offered by 44 of 90 schools (48.9%).

### Evaluation of program effectiveness

Forty-three of 90 schools (47.8%) formally evaluated the effectiveness of their wellness programs; 31 (34.4%) did not, and 16 (17.8%) were unsure. Among the 42 schools that described their methods, AAMC national surveys predominated (39/42, 92.9%), followed by wellness-specific satisfaction ratings (35/42, 83.3%), attendance or participation rates (29/42, 69.0%), and qualitative feedback (27/42, 64.3%). Standardized outcome measurement was uncommon: 11 of 42 schools (26.2%) reported longitudinal tracking with standardized instruments, and 10 (23.8%) reported standardized pre/post assessment, with 14 (33.3%) using either. Reported barriers to evaluation were limited staffing or personnel time (45/84, 53.6%), low student participation (36/84, 42.9%), insufficient finances (29/84, 34.5%), and lack of suitable instruments (22/84, 26.2%). Asked in open text to name evaluation data their institution lacked, 52 schools gave a substantive answer; standardized measures were the most frequently identified need (20/52, 38.5%), followed by longitudinal or repeated measurement (14/52, 26.9%).

### Use and reporting of wellness data

Sixteen of 89 schools (18.0%) systematically analyzed wellness outcomes by student demographic characteristics; 50 (56.2%) did not, and 23 (25.8%) were unsure. No school reported publishing summary wellness outcomes in full; 9 of 84 (10.7%) reported partial publication, 57 (67.9%) reported none, and 18 (21.4%) were unsure.

### The sequence from aims to evidence

Among the 84 schools answering all five linked items, 2 (2.4%) reported all five practices, 22 (26.2%) reported none, and 46 (54.8%) reported no more than one. Twenty (23.8%) reported explicit wellness competencies, 44 (52.4%) reported formal assessment of student well-being, 39 (46.4%) reported formal program evaluation, 14 (16.7%) reported analysis by student demographic characteristics, and 9 (10.7%) reported at least partial publication of summary outcomes (Figure 1). Assessment and evaluation did not consistently occur together: 14 schools reported evaluation without assessment, 19 reported assessment without evaluation, and 26 reported neither.

**Figure 1.**
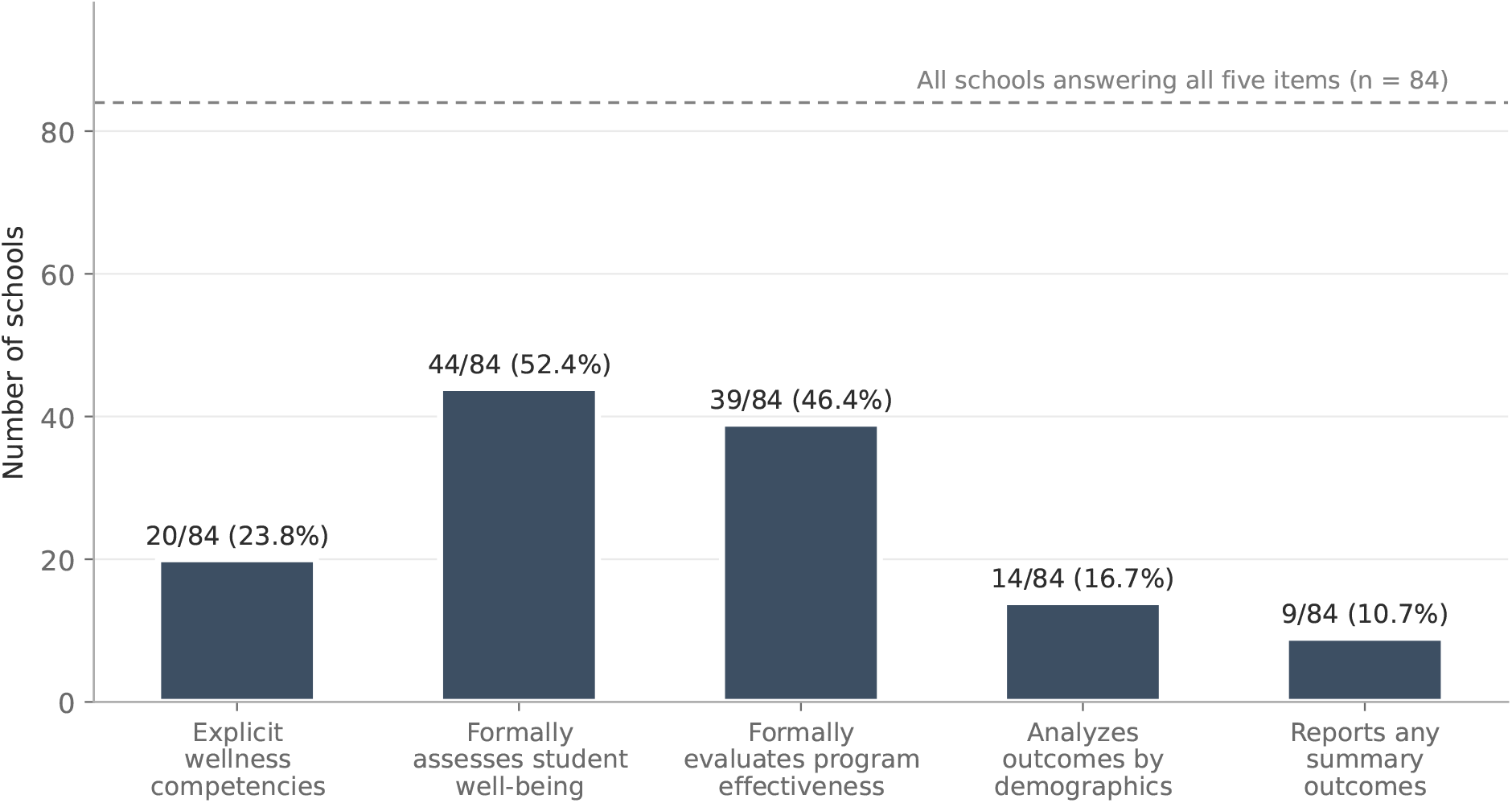
Institutional practices linking stated aims to reported evidence for student wellness programs among 84 U.S. MD-granting medical schools answering all five items, 2025-2026. Bars show the number and percentage of schools reporting each practice; the dashed line marks the complete-case denominator. Two schools (2.4%) reported all five practices, and 22 (26.2%) reported none.

In exploratory analyses, the 20 schools reporting explicit wellness competencies reported more of the four remaining practices than the 64 schools reporting none or unsure (median 2 vs 1; mean 1.9 vs 1.1; *P* = .007). This difference persisted after excluding the 34 unsure schools (*P* = .01). Associations were strongest for program evaluation and outcome reporting: schools with explicit competencies more often reported formal program evaluation (14/20 [70.0%] vs 25/64 [39.1%]; odds ratio [OR] 3.58; 95% CI, 1.11-12.96) and reporting of outcomes (6/20 [30.0%] vs 3/64 [4.7%]; OR 8.41; 95% CI, 1.57-58.37). Formal assessment of student well-being (11/20 [55.0%] vs 33/64 [51.6%]; OR 1.15) and designated wellness leadership (16/20 [80.0%] vs 49/64 [76.6%]; OR 1.22) did not differ by competency status. After Holm adjustment across seven comparisons, only the association with outcome reporting remained statistically significant (adjusted *P* = .03). Excluding unsure schools attenuated associations with demographic analysis and standardized instrument use; associations with program evaluation and outcome reporting remained statistically significant before adjustment (Supplemental Digital Appendix 2).

## Discussion

In this national survey of U.S. MD-granting medical schools, wellness programming and the organizational structures supporting it were widespread, consistent with the two prior multi-institutional surveys.^9,10^ What was not widespread was the capacity to know whether any of it works. Roughly one quarter of schools defined what their wellness efforts are meant to achieve; about half measured student well-being or evaluated programs; fewer than one in five examined outcomes across student groups; and none reported results publicly in full. Two of 84 schools reported the complete sequence the CDC framework describes. The binding constraint in medical student wellness is no longer the absence of programs. Instead, programs are being run without the information needed to manage them.

Prior work has documented individual weak points, particularly reliance on participation and satisfaction measures.^9,10^ Examining the practices together shows that they are not merely each imperfect but largely disconnected from one another. Fourteen schools reported evaluating the effectiveness of their wellness programs while reporting no formal assessment of student well-being, which means program effectiveness was being judged against something other than student outcomes. Nineteen schools measured student well-being without evaluating programs, generating data that informed no stated decision.

The sequence is weakest at its origin. Explicit wellness competencies were reported by 25.3% of schools, essentially unchanged from the 22% Dyrbye and colleagues reported from a much smaller sample nearly a decade earlier.^9^ More striking is that 41.8% of respondents, most of whom hold institutional responsibility for student wellness, did not know whether their school had such competencies. Schools that stated competencies more often evaluated their programs and reported what they found, but were no more likely to have designated wellness leadership or to measure student well-being. The data cannot establish direction, and some of the association may reflect respondents who were better informed about every practice; that the evaluation and reporting associations held when unsure respondents were excluded, while weaker associations did not, argues against informant knowledge as the whole explanation.

Competencies must also be framed carefully; defining a particular state of personal well-being as a standard students must meet would hold learners accountable for remaining well under conditions they do not control.^18,19^ The useful function of stated aims here is institutional: to specify what a school is trying to change so that it becomes possible to ask whether it changed.

Measurement then defaults to what is already available. More than nine in ten schools that assessed student well-being used AAMC national surveys, as did a similar share of those that evaluated programs. These instruments allow comparison with national patterns without additional collection burden, but the AAMC working group has noted their limited wellness-specific content and recommended pairing them with brief standardized measures aligned with local goals.^11^ Over one third of assessing schools used AAMC surveys without any standardized instrument. Half used a standardized general well-being instrument; about one quarter used instruments for burnout, depression, or anxiety. The gap is less about instrument availability than about using instruments in a design that can detect change: only a third of evaluating schools used standardized longitudinal or pre/post measurement.

What happens after measurement deserves separate attention. Among schools that formally assess student well-being, half offered only opt-in support, and nearly one in five offered no follow-up support at all. Assessment that identifies distress without a defined institutional response puts the burden on the student to act on the finding, the opposite of the intent. A school unable to mount longitudinal standardized measurement can still specify what happens when an assessment flags a student.

The final two practices, analyzing outcomes across student groups and reporting findings, were the least common. Fewer than one in five schools examined outcomes by student demographic characteristics, making it difficult to detect differences in well-being, access to support, or program effects where sample size and privacy protections would permit examination. No school reported publishing summary wellness outcomes in full. Unrestricted disclosure of sensitive data is not appropriate, but the practical intermediate step, returning deidentified aggregate findings to students and institutional groups along with an account of what changed as a result, was also rare.

A related mismatch appeared between where schools direct wellness efforts and where their own respondents locate stress. Curricular workload, clerkships, and board examinations were the most frequently named stressors, yet wellness curricula were concentrated in orientation programming, with integration into coursework or rotations less common. Integration competes for time schools named as their main barrier, so the question is less about volume than timing relative to clerkships and examinations. Given the prominence of clerkships among these stressors, the clinical mix also warrants attention: near-universal mistreatment policies and common work-hour protections coexisted with faculty and resident training at fewer than half of schools and flexible scheduling at one third. Whether students experience these policies depends substantially on supervisors, and prospective work has linked mistreatment and poor role modeling during clinical rotations to greater depression and stress symptoms.^20,21^ Supervisor development is therefore a condition of structural protections operating as intended.

Limited staffing, low student participation, and insufficient finances were the leading barriers to evaluation, and respondents most often named standardized measures as the evaluation data they lacked. National organizations are better positioned than individual schools to solve these problems, for example, by curating a concise set of measures with strong validity evidence matched to common wellness aims and publishing adaptable evaluation protocols, extending the direction the AAMC working group has already set.^11^ At the institutional level, the more immediate step is smaller: state what a major wellness initiative is intended to accomplish, select one measure capable of detecting it, define what happens when the measure identifies a student in difficulty, and report back what was found. Most schools in our sample currently complete at most one of these steps.

## Limitations

Each response reflected a single key informant, who may have differed from others in knowledge or interpretation, and self-report may overestimate implementation; student-reported experiences were not available for corroboration. The 67.3% institutional response rate permits nonresponse bias; regional differences did not reach statistical significance, but the observed spread from 57.1% to 80.0% means they cannot be excluded, and the accreditation-status comparison involved too few schools to be informative. Partial responses produced declining denominators; we reported item-specific denominators, did not impute, and found no evidence that completers differed from partial responders, although this comparison is limited to early items. Validity evidence was limited to content and response-process evidence from physician well-being experts who are coauthors, not student-affairs leaders like most respondents. Data were collected across two periods more than a year apart and are combined throughout; the supplementary round contributed 16 schools, too few for a meaningful comparison between rounds. The sequence analysis describes co-occurrence of self-reported practices rather than causal dependency, and the competency analyses were exploratory, based on 20 exposed schools, and yielded wide confidence intervals; they should be treated as hypothesis-generating. The cross-sectional design cannot establish change since prior surveys, which used different sampling frames and item wording.

## Conclusions

U.S. MD-granting medical schools have built substantial student wellness infrastructure, including curricula, learning communities, dedicated leadership, funded committees, and extensive student participation. Far fewer have defined what these efforts are meant to accomplish, measured student well-being with instruments capable of detecting change, evaluated programs against those measures, examined effects across student groups, or reported what they found. Two of 84 schools reported doing all five, and schools that stated their aims were more likely to evaluate and report. Closing this gap is less about adding programs than about making existing ones answerable, starting with stated aims, a measure aligned to them, and a defined response when that measure identifies a student in difficulty.

## Supporting information

Supplemental Digital Appendices 1-2 (survey instrument and exploratory analyses)

## Data Availability

Institution-level data are not publicly available because respondents were assured school-level confidentiality and the data may permit identification of schools. Aggregate data tables and analytic code may be available from the corresponding author on reasonable request, subject to institutional review. The full survey instrument is included in the supplementary material.

## Declarations

### Ethical approval

On March 26, 2025, the Stanford University Institutional Review Board determined that protocol 79558 did not involve human subjects as defined in 45 CFR 46.102(e) or 21 CFR 50.3(g); no further action was required.

### Previous presentations

Preliminary findings from this study were presented orally at the Health Professions Education Conference in Honolulu, Hawaii, on January 31, 2026.

### Data availability

Institution-level data are not publicly available because respondents were assured school-level confidentiality and the data may permit identification of schools. Aggregate data tables and analytic code may be available from the corresponding author on reasonable request, subject to institutional review.

### Author contributions

J.G.: conceptualization, methodology, investigation, data curation, formal analysis, project administration, writing (original draft). M.W.: methodology, writing (review and editing). A.A.: methodology, writing (review and editing). C.P.: conceptualization, methodology, formal analysis, supervision, writing (review and editing).

## Acknowledgments

The authors used Claude (Anthropic, San Francisco, California) to assist with writing analysis code and editing manuscript text. The authors reviewed and verified all code, results, and text and take full responsibility for the content.

