## Supplemental Digital Appendices 1-2 (survey instrument and exploratory analyses) for "Widespread Programs, Limited Evidence: How U.S. MD-Granting Medical Schools Define, Measure, and Evaluate Student Well-Being Efforts"

### Supplemental Digital Appendix 1: Survey Instrument

> *Thank you for participating in our study. Your participation will help us understand and improve student wellness practices across medical schools. Individual responses at the school level will be kept confidential and anonymous. This survey should take about 15 to 20 minutes to complete.*

#### Section 1. Curriculum and Learning Environment

*This first block of questions focuses on curricular interventions and the learning environment at your school.*

**Q1.** Does your school have a dedicated wellness curriculum (i.e., structured programming integrated into the formal academic curriculum to promote student well-being)?

○ Yes

○ No

○ No, but in development or actively planning

**Q2.** How is the wellness curriculum structured? (Select all that apply.)

*Displayed if Q1 response was not "No".*

☐ Integrated into existing academic coursework or rotations

☐ Standalone courses or workshops

☐ Part of orientation programming

☐ Other (please specify) ________________

☐ Unsure

**Q3.** Who is targeted by your wellness curriculum? (Select all that apply.)

*Displayed if Q1 response was not "No".*

☐ Preclinical students

☐ Clinical students

☐ Other (please specify) ________________

☐ Unsure

**Q4.** How are clinical rotations organized at your institution?

○ Block rotations

○ Longitudinal integrated clerkship

○ Other (please specify) ________________

○ Unsure

**Q5.** Which wellness-oriented interventions or policies are incorporated into clinical rotations at your institution? (Select all that apply.)

☐ Work-hour limitations or protected time off (e.g., "wellness days")

☐ Wellness/reflection check-ins with mentors or faculty

☐ Flexible scheduling that accommodates students' preferences and personal circumstances

☐ Explicit avenues/policies to report and prevent mistreatment

☐ Training/guidance for faculty and residents on promoting wellness

☐ Provision of healthy food and/or guaranteed meal breaks

☐ Grading or assessment modifications that reduce stress (e.g., pass/fail evaluations, standardized rubrics)

☐ Other (please specify) ________________

**Q6.** Does your institution offer training or professional development for faculty aimed at promoting a wellness-friendly environment? (Select all that apply.)

☐ Training/workshops designed to enhance faculty well-being (e.g., stress management, work–life balance)

☐ Training/workshops designed to help faculty support student wellness (e.g., identifying burnout)

☐ Other (please specify) ________________

☐ No

☐ Unsure

**Q7.** Does your medical school offer structured learning communities (e.g., longitudinal societies, colleges, houses, mentorship families/cohorts)?

○ Yes

○ No

○ Unsure

**Q8.** Please indicate for each component below whether it is integrated into these learning communities.

*Displayed if Q7 response was "Yes".*

| | Yes | No | Not sure |
|---|---|---|---|
| Inclusion of Students from Different Years (e.g., cross-year interactions, pairing upperclassmen with junior students) | ○ | ○ | ○ |
| Inclusion of Faculty (e.g., regular faculty engagement or mentoring) | ○ | ○ | ○ |
| Structured Social Events and Community-Building Activities | ○ | ○ | ○ |
| Academic Advising and Support | ○ | ○ | ○ |
| Clinical/Doctoring Skills Development (e.g., practicing physical examination techniques) | ○ | ○ | ○ |
| Career Planning and Professional Development | ○ | ○ | ○ |
| Wellness, Self-Care, & Resilience Programming | ○ | ○ | ○ |
| Research or Quality Improvement Projects | ○ | ○ | ○ |
| Community Service & Outreach | ○ | ○ | ○ |

**Q9.** We welcome you to use this space to describe any notable or exemplary curricular strategies your school uses to promote medical student wellness (e.g., longitudinal electives, innovative grading systems, small-group learning):

______________________________

**Q10.** Which elements of your academic curriculum (e.g., a particular course or clinical rotation) create the greatest stress or demands on students? List up to three.

1. ______________________________
2. ______________________________
3. ______________________________

**Q11.** What are the primary barriers your institution faces in implementing curricular or structural changes designed to improve medical student wellness? (Select all that apply)

☐ Lack of financial resources

☐ Limited administrative or leadership support

☐ Faculty resistance or lack of buy-in

☐ Curricular overcrowding (e.g., insufficient time or space in the curriculum to integrate wellness programming)

☐ Logistical/scheduling difficulties

☐ Other (please specify) _______________

### Section 2. Assessment and Evaluation of Wellness

*Thanks for your responses so far! In the next block of questions, we'd like to learn about how your medical school evaluates student well-being and the effectiveness of wellness programming.*

**Q12.** Are any competencies (i.e., milestones, entrustable professional activities, learning outcomes, etc.) in self-care or physician wellness explicitly stated in your institution's program objectives or formal competency framework?

○ Yes (please list them or send documents to [email address redacted for publication]) _______________

○ No

○ Unsure

**Q13.** Does your school formally assess medical student wellness?

○ Yes

○ No

○ Unsure

**Q14.** Which methods does your institution use to assess student wellness? (Select all that apply.)

*Displayed if Q13 response was "Yes".*

☐ Information from AAMC national surveys (e.g., Year Two Questionnaire [Y2Q], Graduation Questionnaire [GQ])

☐ Confidential clinical evaluations

☐ Attitudinal/experience-based student wellness surveys

☐ Focus groups, town hall meetings, or qualitative interviews

☐ Standardized, validated instruments for burnout (e.g., Maslach Burnout Inventory)

☐ Standardized, validated instruments for depression (e.g., PHQ-9)

☐ Standardized, validated instruments for anxiety (e.g., GAD-7)

☐ Other standardized, validated instruments for well-being (e.g., Medical Student Well-Being Index, Professional Quality of Life Scale, Perceived Stress Survey)

☐ Other (please specify) ______________

**Q15.** For each applicable method selected, please indicate the frequency of administration (e.g., annually, semesterly, quarterly) if known:

*Displayed if Q13 response was "Yes".*

______________________

**Q16.** Does your institution provide any follow-up or support based on results from the screening or assessment methods mentioned above? If yes, please use the accompanying text box to elaborate on the result thresholds or criteria that trigger each type of support.

*Displayed if Q13 response was "Yes".*

○ Yes, opt-in support available (students are offered services but must request them) ______________

○ Yes, opt-out support provided (students meeting defined criteria are automatically enrolled unless they decline) ______________

○ No

○ Unsure

**Q17.** Does your institution offer any anonymous, self-administered screening tools to help students assess their own wellbeing or mental health over time?

○ Yes

○ No

○ Unsure

**Q18.** Please briefly describe the anonymous self-screening tools (e.g., web page linking to PHQ-9, third-party digital platform, custom in-house software). If possible, include details on whether any follow-up or support is provided based on screening results:

*Displayed if Q17 response was "Yes".*

______________________

**Q19.** Does your institution systematically collect and analyze demographic data (year, race/ethnicity, gender) in relation to wellness outcomes, such that differences/disparities may be identified across different groups?

○ Yes (describe any disparities identified and actions taken): ______________

○ No

○ Unsure

**Q20.** Does your school formally evaluate the effectiveness of its wellness program? (For this survey, a wellness program is defined as an organized, institutionally supported set of practices or initiatives designed to enhance medical students' mental, emotional, physical, social, and professional well-being.)

○ Yes

○ No

○ Unsure

**Q21.** Which methods does your institution use to evaluate its wellness program? (Select all that apply)

*Displayed if Q20 response was "Yes".*

□ Information from AAMC national surveys (e.g., Year Two Questionnaire [Y2Q], Graduation Questionnaire [GQ])

□ Student satisfaction ratings from wellness-specific surveys or course evaluations

□ Qualitative feedback from focus groups, town hall meetings, interviews or open-ended survey questions

□ Attendance/participation rates in wellness programs

□ Pre-/post-intervention assessments using validated instruments (e.g., Maslach Burnout Inventory, Medical Student Well-Being Index)

□ Longitudinal tracking of wellness/burnout/anxiety/depression using validated instruments

□ Faculty feedback/ratings regarding wellness offerings

□ Other (please specify): ________________

**Q22.** For each applicable method selected above, please indicate how frequently it is administered (e.g., annually, semesterly, quarterly), if known:

*Displayed if Q20 response was "Yes".*

________________________________

**Q23.** We welcome you to use this space to describe the process and/or frequency with which your school reviews evaluation data and uses it to inform program modifications:

*Displayed if Q20 response was "Yes".*

________________________________

**Q24.** Please identify at least one type of data or evaluation method that your institution does not currently collect/utilize, but which would significantly enhance the development of your wellness programming if implemented:

________________________________

**Q25.** Considering the gap(s) you've identified in the previous question, as well as your institution's wellness program evaluation practices more generally, what are the primary barriers your school faces in evaluating its wellness initiatives? (Select up to 3.)

□ Lack of suitable instruments/surveys

□ Low student participation or engagement

□ Faculty resistance or low faculty buy-in

□ Limited staffing or personnel time

☐ Insufficient financial resources

☐ Lack of administrative or leadership support

☐ Lack of expertise

☐ Other (please specify) ________________

**Q26.** Does your institution publish any summary data or evaluation reports on wellness outcomes for public review?

○ Yes (please specify or provide a link) ________________

○ No

○ Partially

○ Unsure

### Section 3. Organizational and Operational Resources

*Thanks again for your continued input. You're doing great! The following set of questions will address how your medical school organizes and operates its wellness initiatives.*

**Q27.** Does your school have a student wellness committee?

○ Yes – Student-only or mostly students

○ Yes – Student and faculty collaboration

○ Yes – Faculty-only or mostly faculty

○ No

○ Unsure

**Q28.** What are the main responsibilities of the wellness committee at your institution? (Select all that apply.)

*Displayed if Q27 response was not "No" and Q27 response was not "Unsure".*

☐ Planning and organizing wellness events

☐ Policy advocacy for student wellness

☐ Coordinating peer support and outreach initiatives

☐ Liaising with faculty and administration

☐ Managing a dedicated wellness budget

☐ Other (please specify) ________________

☐ Unsure

**Q29.** Besides wellness committee membership, in what additional ways are students involved in planning or leading wellness initiatives? (Select all that apply.)

☐ Leading student wellness organizations or clubs

☐ Participating in the co-design of wellness curricula and programming

☐ Securing unstructured funding or grant opportunities for wellness projects

☐ Organizing independent peer mentoring or counseling programs

☐ Conducting student-led research and quality improvement projects for wellness

☐ Serving as wellness ambassadors or in other designated leadership roles

☐ Other (please specify) ________________

☐ Unsure

**Q30.** You may use this space to provide details or examples of any particularly successful student-led wellness initiatives.

________________________________

**Q31.** Does your institution have a designated individual in its medical school administration whose primary responsibility is to oversee student wellness?

○ Yes

○ No

○ Unsure

**Q32.** If known, please provide the title, approximate protected time (% FTE), and reporting structure of the individual(s) who oversee medical student wellness at your institution.

*Displayed if Q31 response was "Yes".*

________________________________

**Q33.** What are the primary funding sources for your wellness initiatives? (Select all that apply.)

☐ Dedicated budget line within the institution

☐ External grants

☐ Philanthropic donations

☐ Integrated into the overall institutional budget

☐ Other (please specify) ________________

☐ Unsure

**Q34.** Which digital resources or online tools does your institution offer to support student wellness? (Select all that apply.)

☐ Mental health screening and self-assessment tools (e.g., Well-Being Index)

☐ Digital resource libraries/portals with curated wellness content (videos, podcasts, articles)

☐ Stress management, mindfulness/meditation, or wellness apps

☐ AI-based chatbots for mental health or wellness support

☐ Virtual therapy or telemedicine platforms

☐ Digital appointment scheduling for counseling or wellness services

☐ Virtual peer support platforms or online community forums

☐ Virtual reality (VR) guided relaxation/meditation experiences

☐ Other (please specify) ________________

### Section 4. Additional Information and Future Directions

*We're almost done—thank you! Please use these final questions to offer any additional comments, challenges, or plans for the future of your wellness programming.*

**Q35.** Please list up to three initiatives or aspects of your wellness program that have proven to be the most effective.

1. ______________________________
2. ______________________________
3. ______________________________

**Q36.** We welcome you to share any further challenges, future plans (especially related to the topics covered in this survey), URLs/links, or other information regarding your institution's wellness initiatives.

______________________________

### Supplemental Digital Appendix 2

#### Exploratory Associations Between Explicit Wellness Competencies and Other Institutional Practices Among 84 U.S. MD-Granting Medical Schools Answering All Five Linked Items, 2025-2026

| | Primary comparison | | | | | Sensitivity (unsure excluded) | | |
|---|---|---|---|---|---|---|---|---|
| Practice | Yes (n = 20) | No/unsure (n = 64) | OR (95% CI) | *P* | Holm *P* | No (n = 30) | OR | *P* |
| Formally assesses student well-being | 11 (55.0) | 33 (51.6) | 1.15 (0.37-3.60) | .80 | > .99 | 12 (40.0) | 1.81 | .39 |
| Formally evaluates program effectiveness | 14 (70.0) | 25 (39.1) | 3.58 (1.11-12.96) | .02 | .13 | 11 (36.7) | 3.91 | .04 |
| Uses any standardized well-being instrument | 8 (40.0) | 18 (28.1) | 1.69 (0.51-5.43) | .41 | > .99 | 11 (36.7) | 1.15 | > .99 |
| Analyzes outcomes by demographic characteristics | 7 (35.0) | 7 (10.9) | 4.29 (1.08-17.30) | .03 | .17 | 5 (16.7) | 2.64 | .18 |
| Any reporting of summary outcomes | 6 (30.0) | 3 (4.7) | 8.41 (1.57-58.37) | .005 | .03 | 2 (6.7) | 5.77 | .047 |
| Designated administrative wellness leader | 16 (80.0) | 49 (76.6) | 1.22 (0.32-5.79) | > .99 | > .99 | 23 (76.7) | 1.21 | > .99 |
| Student wellness committee | 19 (95.0) | 54 (84.4) | 3.48 (0.44-160.61) | .45 | > .99 | 24 (80.0) | 4.63 | .22 |

Number of the four remaining sequence practices (formal assessment, formal evaluation, demographic analysis, any reporting) reported: competencies yes, median 2 (mean 1.90); no or unsure, median 1 (mean 1.06); Mann-Whitney $P = .007$. Excluding unsure respondents: no, median 1 (mean 1.00); $P = .01$.

Abbreviations: CI, confidence interval; OR, odds ratio. Odds ratios are conditional maximum likelihood estimates with exact confidence intervals; *P* values are from 2-sided Fisher exact tests. Holm adjustment was applied across the 7 practice-level comparisons. Schools not reporting formal assessment were counted as not using a standardized instrument. Analyses are exploratory and hypothesis-generating.
